# Impact of COVID-19 mitigation measures on the incidence of preterm birth: a national quasi-experimental study

**DOI:** 10.1101/2020.08.01.20160077

**Authors:** Jasper V Been, Lizbeth Burgos Ochoa, Loes CM Bertens, Sam Schoenmakers, Eric AP Steegers, Irwin KM Reiss

## Abstract

**Introduction:** Preterm birth is the leading cause of child mortality globally, with many survivors experiencing long-term adverse consequences. Preliminary evidence suggests that preterm births dropped dramatically following implementation of policy measures aimed at mitigating the impact of the COVID-19 pandemic.

**Methods:** We undertook a national quasi-experimental difference-in-regression-discontinuity approach to study the impact of the COVID-19 mitigation measures implemented in the Netherlands in a stepwise fashion on 9, 15, and 23 March 2020 on the incidence of preterm birth. We used data from the neonatal dried blood spot screening programme (2010−2020) and cross-validated these against national perinatal registry data. Stratified analyses were conducted according to gestational age subgroups, and sensitivity analyses to assess robustness of the findings. We explored potential effect modification by neighbourhood socio-economic status.

**Results:** Data on 1,599,547 singleton newborns were available, including 56,720 post-implementation births. Consistent reductions in preterm birth were seen across various time windows surrounding implementation of the 9 March COVID-19 mitigation measures: ±2 months (n=531,823): odds ratio 0.77 (95% confidence interval 0.66–0.91), p=0.002; ±3 months (n=796,531): 0.85 (0.73–0.98), p=0.028; ±4 months (n=1,066,872): 0.84 (0.73–0.97), p=0.023. Decreases observed following the 15 March measures were of smaller magnitude and not statistically significant. No changes were observed after 23 March. Preterm birth reductions after 9 March were consistent across gestational age strata and robust in sensitivity analyses. They appeared confined to high-socioeconomic status neighbourhoods, but effect modification was not statistically significant.

**Conclusion:** In this national quasi-experimental study, initial implementation of COVID-19 mitigation measures was associated with a 15-23% drop in preterm births in the following months, in agreement with preliminary observations in other countries. It is now of pivotal importance that integration of comparable data from across the globe is undertaken to further substantiate these findings and start exploring the underlying mechanisms.

## Background

The COVID-19 pandemic and the measures taken to help prevent spread of infection and mitigate its population health effects are having an unprecedented impact on society. The sudden occurrence of the pandemic and the scale and immediacy of the policy responses taken, provide a unique opportunity to evaluate their effects as a ‘natural experiment’.^1^ Intriguingly, recent reports from Denmark and Ireland independently provided evidence indicating substantial reductions in the number of extremely preterm and very-low-birth-weight (VLBW) births following national COVID-19 mitigation measures.^2,3^ Several potential underlying mechanisms have been proposed, including improvements in ambient air quality, and reductions in maternal stress and incidence of infections.^3^

Globally, over one in ten babies are born preterm, and preterm birth is the primary contributor to mortality in early life.^4^ In addition, preterm birth survivors and their families frequently experience long-term adverse consequences.^5-8^ Currently, the opportunities for prevention of preterm birth are very limited.^9^ As such, it is of pivotal importance that we further explore the possible link between national lockdown measures and a decrease in preterm births, and if confirmed, start identifying the underlying mechanisms to inform and optimise future approaches to help prevent preterm birth from devastating families’ lives.

At present, although the link between COVID-19 mitigation measures and reductions in preterm birth identified in the pioneering aforementioned Danish and Irish studies has rightfully sparked substantial optimism globally regarding its potential to help identify new clues for effective prevention, the evidence base is still delicate.^2,3^ Both previous studies had relatively limited sample sizes and the methodological approaches that were used restrict causal interpretation of the findings. In the current study we addressed these limitations by using national routinely collected data to study the association between the implementation of COVID-19 mitigation measures in the Netherlands and the incidence of preterm birth. We applied a difference-in-regression-discontinuity design, facilitating casual inference over the non-quasi-experimental approaches used in previous studies.^2,3^

## Methods

We undertook a difference-in-regression-discontinuity analysis to investigate the association between the national implementation of COVID-19 mitigation measures and the incidence of preterm birth, using national routinely collected data on singleton babies having undergone neonatal blood spot screening in the Netherlands between October 2010 and July 2020.

### Setting and participants

The first recognised COVID-19 case in the Netherlands was confirmed in Noord-Brabant, one of twelve Dutch provinces, on 27 February 2020.^10^ The first COVID-19-related death occurred on 6 March, and from that day people living in Noord-Brabant were advised to stay indoors when experiencing possible COVID-19 symptoms. On separate occasions between 9 and 23 March, a number of national measures were then taken and widely communicated in an attempt to mitigate the impact of the COVID-19 pandemic in the Netherlands (Table 1).^10^

**Table 1.** Timeline of implementation of key COVID-19 mitigation measures in the Netherlands.

| Date | COVID-19 mitigation measures implemented |
| --- | --- |
| **9 March 2020** | Strong advice against handshaking, and for using paper handkerchiefs, sneezing/coughing in one's elbow, and regular handwashing<br>Strong advice for staying at home when experiencing cold symptoms or fever, or when having been in contact with COVID-19-positive person or having visited a high-risk area |
| **12 March 2020** | Strong advice against social interaction, and against visiting elderly people<br>Events of >100 individuals are cancelled<br>People need to work from home whenever possible<br>People need to stay home if symptomatic (fever, respiratory complaints) |
| **15 March 2020** | Closing down of schools and child care facilities<br>Closing down of hospitality industry and of non-essential services involving physical contact |
| **23 March 2020** | All events and gatherings are cancelled<br>Physical distancing is introduced (1.5-meter-rule)<br>Issuing of fines for not complying with physical distancing<br>Municipalities may close down busy places and shops |

We obtained data on all singleton babies having undergone neonatal blood spot screening in the Netherlands between 9 October 2010 and 16 July 2020, the latter date representing the most recent data available at the time of extraction. The study period was set to include ten years and five months pre-implementation of the first national COVID-19 mitigation measures (9 March 2020; Table 1). Data were extracted from the Praeventis database, as provided by the National Institute for Public Health and the Environment (RIVM).^11^ Praeventis is a national database containing data from all babies having undergone neonatal blood spot screening. In the national screening programme, newborns are screened for a range of diseases after 72 hours of life. Screening can take place in the hospital or at home. The proportion of Dutch babies undergoing neonatal blood spot screening is consistently >99%,^12^ hence the Praeventis database may be considered to be highly representative of all births in the Netherlands. On the neonatal dried blood spot card, health professionals record several maternal and neonatal characteristics for each registered individual.^13^

For the purpose of this study, multiple births were excluded due to their inherent increased risk of preterm birth. Multiple births were identified based on having multiple records registered with identical surnames, birth dates and postcode. We furthermore excluded babies whose registered gestational age was below 24+0 weeks or above 41+6 weeks. Dutch national multidisciplinary guidelines have set the threshold of viability at 24+0 weeks and advise against active management of babies born at lower gestational ages.^14^

For validation purposes, characteristics of our cohort were cross-referenced at aggregate level against data from Perined for selected years. Perined is the national linked pregnancy and birth registry which is based on data provided by midwifery, general practice, and obstetric and paediatric practices.^15^ Perined data are typically made available 1-2 years following initial registration of pregnancies and births, hence invalidating the use of Perined data to address our primary research question at present.

### Variables and data source

The following individual-level data were extracted from Praeventis: 1. calendar week of birth; 2. gestational age (in days); 3. birth weight (in grams); 4. sex; and 5. four-digit postcode. Four-digit postcode identifies areas with an average of 2,160 households and was used to derive: 1. province of residence; 2. neighbourhood socioeconomic status (SES; in quintiles); and 3. neighbourhood urbanisation level. Neighbourhood SES scores are calculated by The Netherlands Institute for Social Research (SCP) since 1998 and updated every four years since.^16^ SES scores are based on: mean household income, proportion of population with low income, proportion of population with low educational level, proportion of population without paid work. Urbanisation level was dichotomised, with urban areas having >2,500 addresses per km^2^. Individual-level sex- and gestational age-specific birth weight centiles were calculated using national reference curves.^17^

### Sample size

Two earlier studies have identified a link between national implementation of COVID-19 mitigation measures and a reduction in extremely preterm and VLBW births.^2,3^ In the Danish study,^2^ national data on post-implementation births were available for 5,162 singleton births. The Irish study was a single-centre study and had 1,381 births available for analysis.^3^ The Netherlands has approximately 170,000 births annually. This translates into an anticipated ∼60,000 births post-implementation, including ∼4,000 preterm births. We anticipated that this would provide ample statistical power to identify an association between the COVID-19 mitigation measures and preterm births in the Netherlands, if present.

### Statistical analyses

We tabulated characteristics of the study population according to the time periods from which they were derived. We furthermore tabulated selected characteristics against published Perined annual reports, available up to 2018.^18^

We studied the association between national implementation of the COVID-19 mitigation measures and the incidence of preterm births using a difference-in-regression-discontinuity approach.^19,20^ Difference-in-regression-discontinuity is a quasi-experimental technique that can be used when the exposure of interest is assigned by the value of a continuously measured random variable and whether that variable lies above (or below) some cut-off value. In this study, calendar week of birth is the assignment variable and the cut-off corresponds to the implementation dates of COVID-19 mitigation measures. We conducted separate analyses for the 9, 15, and 23 March implementation of COVID-19 mitigation measures (Table 1). A separate analysis was not possible for the 12 March measures given temporal granularity of the individual-level data (i.e. weekly rather than daily). We *a priori* hypothesised that any reductions in preterm birth would most likely have followed the 15 March 2020 measures as these were considered to be most comprehensive. We assessed four time-windows before and after the intervention in separate analyses: one, two, three, and four months pre- and post-implementation. Using such relatively short discrete time allows us to exclude other interventions or major influences, and make the assumption that any change observed is indeed due to the COVID-19 mitigation measures. The approach allows for comparison of the incidence of preterm birth in the period directly preceding implementation of the measures versus the period directly following implementation. With the shortest time window (i.e. one month), the estimated impact of the implementation of COVID-19 mitigation measures may be closest to the true immediate impact, but the power to detect this impact is limited. Using wider time windows, power to detect the true impact will increase, but potentially at the expense of introducing variation from temporal trends or unmeasured confounding. The analyses account for seasonal variation and potential other time-variant factors affecting preterm birth incidence by comparing the period surrounding the implementation of the measures in 2020 to the exact same time periods in each year preceding the COVID-19 pandemic (2010-2019). By following this approach there is no need to adjust for individual-level variables in the analysis.

The assumptions and conditions for a valid regression discontinuity were met: a) the cut-off value (9, 15 or 23 March 2020) and decision rule (exposed or unexposed to COVID-19 mitigation measures) are known; b) the assignment variable (week of birth) is continuous around the cut-off and not affected by the lockdown, as shown in the Supplement page 1; c) the outcomes are continuous at the threshold and are observed for all pregnancies; d) graphical analysis shows a discontinuity around the threshold, suggesting an intervention effect (Supplement page 2-13).

In the primary analyses, the outcome of interest was the overall incidence of preterm birth (i.e. number of babies born at a gestational age <37+0 weeks per 1,000 babies having undergone neonatal blood spot screening). In additional stratified analyses we assessed whether there were any differential changes in preterm birth incidence following the COVID-19 mitigation measures according to the degree of prematurity: 24+0 – 25+6 weeks, 26+0 – 27+6 weeks, 28+0 – 31+6 weeks, and 32+0 – 36+6 weeks.

Substantial evidence indicates that the COVID-19 pandemic and the measures taken to mitigate its impact are differentially affecting socio-economic groups.^21,22^ To assess whether there has been variation in impact of the Dutch COVID-19 mitigation measures according to SES, we tested for potential effect modification by neighbourhood SES by including an interaction term in the analyses.

Some mechanisms potentially underlying a link between the COVID-19 mitigation measures and preterm birth may not have an immediate impact. On the other hand, there may have been anticipatory effects as part of the population may already have changed their behavior prior to formal implementation of the COVID-19 mitigation measures. To address this we conducted two sets of sensitivity analyses introducing a period of censoring of data, thus excluding data from the first week and from the first two weeks directly prior to and directly following introduction of the measures.

### Ethical considerations

According to Dutch law (WMO) no formal ethical review was required for this study. According to standard procedures and under strict conditions that were fulfilled, RIVM allows anonymised data registered as part of the screening programme to be used for research purposes with waiver of consent.^23^ A protocol for the study was develop *a priori* and approved by RIVM prior to data provision.

## Results

There were 1,707,594 records in the Praeventis neonatal screening database in the study period. After exclusion of neonates born outside the Netherlands, duplicate records, multiple births, and neonates with gestational age missing, <24+0 weeks or >41+6 weeks, individual-level data on 1,599,547 singleton neonates were available for analysis (Figure 1). Characteristics of this population are shown in Table 2. Cross-validation against Perined data for selected years (i.e. 2011, 2014 and 2017) showed that babies born at the lowest gestational ages and those with the lowest birth weights were somewhat underrepresented in our population, which was stable over time (Supplement page 14).

**Table 2:** Characteristics of the study population.

| Characteristic | n (%) / Mean (SD) |
| --- | --- |
| **Birth characteristics** |  |
| Preterm birth (<37+0 weeks) | 84,209 (5.2) |
| 32+0 – 36+6 weeks | 72,753 (4.5) |
| 28+0 – 31+6 weeks | 8,248 (0.5) |
| 26+0 – 27+6 weeks | 2,114 (0.1) |
| 24+0 – 25+6 weeks | 1,094 (0.1) |
| Gestational age (weeks) | 39.5 (1.7) |
| Birthweight (grams)<sup>a</sup> | 3,436 (547) |
| Birthweight centile<sup>a</sup> | 49.3 (29.3) |
| Small for gestational age<sup>a</sup> | 171,910 (10.7) |
| **Sex<sup>b</sup>** |  |
| Male | 819,886 (51.2) |
| Female | 779,654 (48.8) |
| **Province of residence<sup>c</sup>** |  |
| Drenthe | 39,344 (2.5) |
| Flevoland | 45,072 (2.8) |
| Friesland | 57,112 (3.6) |
| Gelderland | 181,830 (11.4) |
| Groningen | 49,643 (3.1) |
| Limburg | 82,613 (5.2) |
| Noord-Brabant | 221,212 (13.8) |
| Noord-Holland | 273,616 (17.1) |
| Overijssel | 109,762 (6.9) |
| Utrecht | 137,630 (8.6) |
| Zeeland | 31,278 (1.9) |
| Zuid-Holland | 369,084 (23.1) |
| **Living in urban area<sup>c</sup>** | 590,028 (36.9) |
| **Neighbourhood socio-economic status<sup>d</sup>** |  |
| Low (<p20) | 301,611 (18.8) |
| Medium (p20-80) | 970,522 (60.7) |
| High (≥p80) | 319,809 (20.0) |

SD = standard deviation; <sup>a</sup>Birth weight was missing for 391 individuals (0.02%); <sup>c</sup>Sex was unspecified for <10 individuals – according to RIVM policy, cells containing <10 individuals are censored; <sup>c</sup>Postcode was missing for 1,195 individuals (0.07%); <sup>d</sup>7,605 cases (0.5%) could not be assigned to an SCP SES category: 1,195 due to missing postcode, 6,410 because SCP does not calculate neighbourhood SES scores for postcodes with less than 100 households

**Figure 1:**
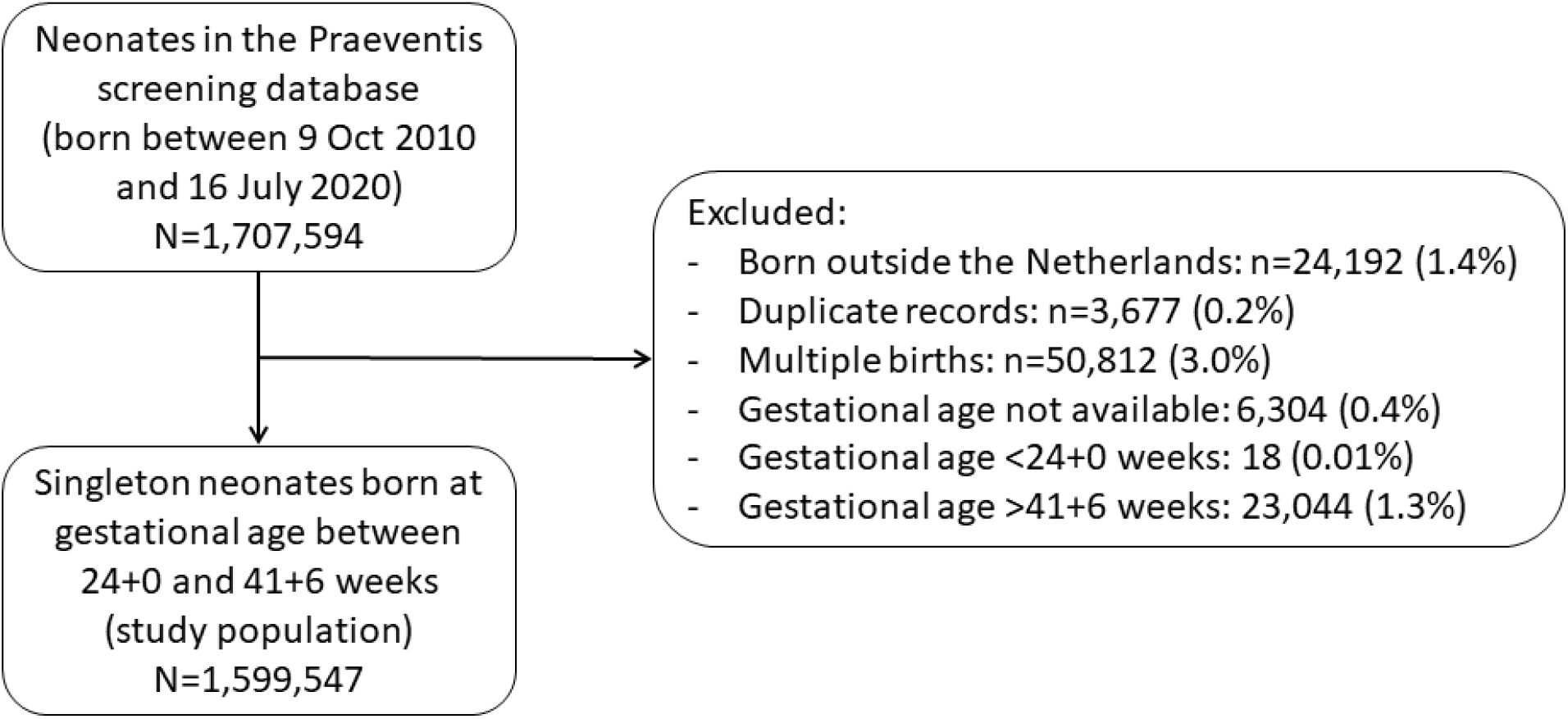
Flowchart of study population composition.

Time trends in preterm births in the four months pre- and post-implementation of the COVID-19 mitigation measures are shown in Figure 2 and Supplement page 2-13. A clear discontinuity in the regression lines is observed when considering the initial set of COVID-19 mitigation measures introduced on 9 March 2020. Accordingly, implementation of the 9 March measures was consistently associated with substantial reductions in preterm birth across the two-to four-month time windows surrounding implementation: ±2 months (n=531,823): odds ratio (OR) 0.77 (95% confidence interval (CI) 0.66–0.91), p=0.002; ±3 months (n=796,531): 0.85 (0.73–0.98), p=0.03; ±4 months (n=1,066,872): 0.84 (0.73–0.97), p=0.02 (Table 3). These reductions in preterm births were apparent across gestational age strata, albeit statistically significant only in the 32+0 to 36+6-week subgroup (Table 3). No significant impact on preterm birth was observed when considering the dates that the initial 9 March measures were extended as the primary intervention dates (i.e. 15 and 23 March; Table 3).

**Table 3:** Impact of COVID-19 mitigation measures on the incidence of preterm birth.

| Intervention / Outcome | Time window around implementation |  |  |  |
| --- | --- | --- | --- | --- |
|  | ±1 month | ±2 months | ±3 months | ±4 months |
| **Measures introduced on 9 March** | **n=262,600** | **n=531,823** | **n=796,531** | **n=1,066,872** |
| Preterm birth (<37+0 weeks) | 0.91 (0.68–1.20) | 0.77(0.66–0.91) | 0.85 (0.73–0.98) | 0.84 (0.73–0.97) |
| 32+0 – 36+6 weeks | 0.91 (0.67–1.23) | 0.78(0.66–0.94) | 0.85 (0.72–0.99) | 0.83 (0.71–0.97) |
| 28+0 – 31+6 weeks | 0.80 (0.34–1.89) | 0.78(0.46–1.33) | 0.88 (0.55–1.40) | 0.91 (0.58–1.42) |
| 26+0 – 27+6 weeks | 1.57 (0.20–12.00) | 0.66(0.21–2.05) | 0.82 (0.30–2.21) | 0.99 (0.38–2.55) |
| 24+0 – 25+6 weeks | 0.89 (0.10–13.00) | 0.48(0.13–1.76) | 0.90 (0.29–2.81) | 1.00 (0.33–3.04) |
| **Measures introduced on 15 March** | **n=259,825** | **n=528,464** | **n=797,799** | **n=1,065,261** |
| Preterm birth (<37+0 weeks) | 1.17 (0.91–1.49) | 0.96 (0.81–1.13) | 0.97 (0.84–1.13) | 0.96 (0.83–1.10) |
| 32+0 – 36+6 weeks | 1.11 (0.58–1.45) | 0.95 (0.79–1.13) | 0.95 (0.82–1.11) | 0.92 (0.80–1.07) |
| 28+0 – 31+6 weeks | 1.30 (0.48–2.23) | 0.88 (0.51–1.50) | 0.96 (0.61–1.51) | 1.00 (0.65–1.55) |
| 26+0 – 27+6 weeks | 4.96 (0.68–36.05) | 1.33 (0.41–4.28) | 1.37 (0.50–3.69) | 1.60 (0.62–4.13) |
| 24+0 – 25+6 weeks | 7.83 (0.73–83.47) | 1.89 (0.48–7.29) | 2.03 (0.63–6.50) | 2.15 (0.69–6.68) |
| **Measures introduced on 23 March** | **n=263,098** | **n=531,720** | **n=799,511** | **n=1,067,665** |
| Preterm birth (<37+0 weeks) | 1.27(0.99–1.60) | 1.06(0.89–1.25) | 1.05(0.91–1.22) | 1.03(0.90–1.18) |
| 32+0 – 36+6 weeks | 1.27(0.99–1.64) | 1.07(0.90–1.28) | 1.05(0.90–1.22) | 1.01(0.87–1.17) |
| 28+0 – 31+6 weeks | 1.18(0.56–2.48) | 0.98(0.57–1.67) | 1.08(0.69–1.69) | 1.12(0.73–1.72) |
| 26+0 – 27+6 weeks | 1.26(0.22–7.09) | 0.89(0.28–2.83) | 1.10(0.42–2.87) | 1.33(0.54–3.29) |
| 24+0 – 25+6 weeks | 0.45(0.07–3.06) | 0.92(0.26–3.26) | 1.22(0.42–3.55) | 1.31(0.46–3.68) |

Odds ratios (95% confidence intervals) from difference-in-regression-discontinuity analysis of various time windows around national implementation of COVID-19 mitigation measures versus the same periods across previous years.

**Figure 2:**
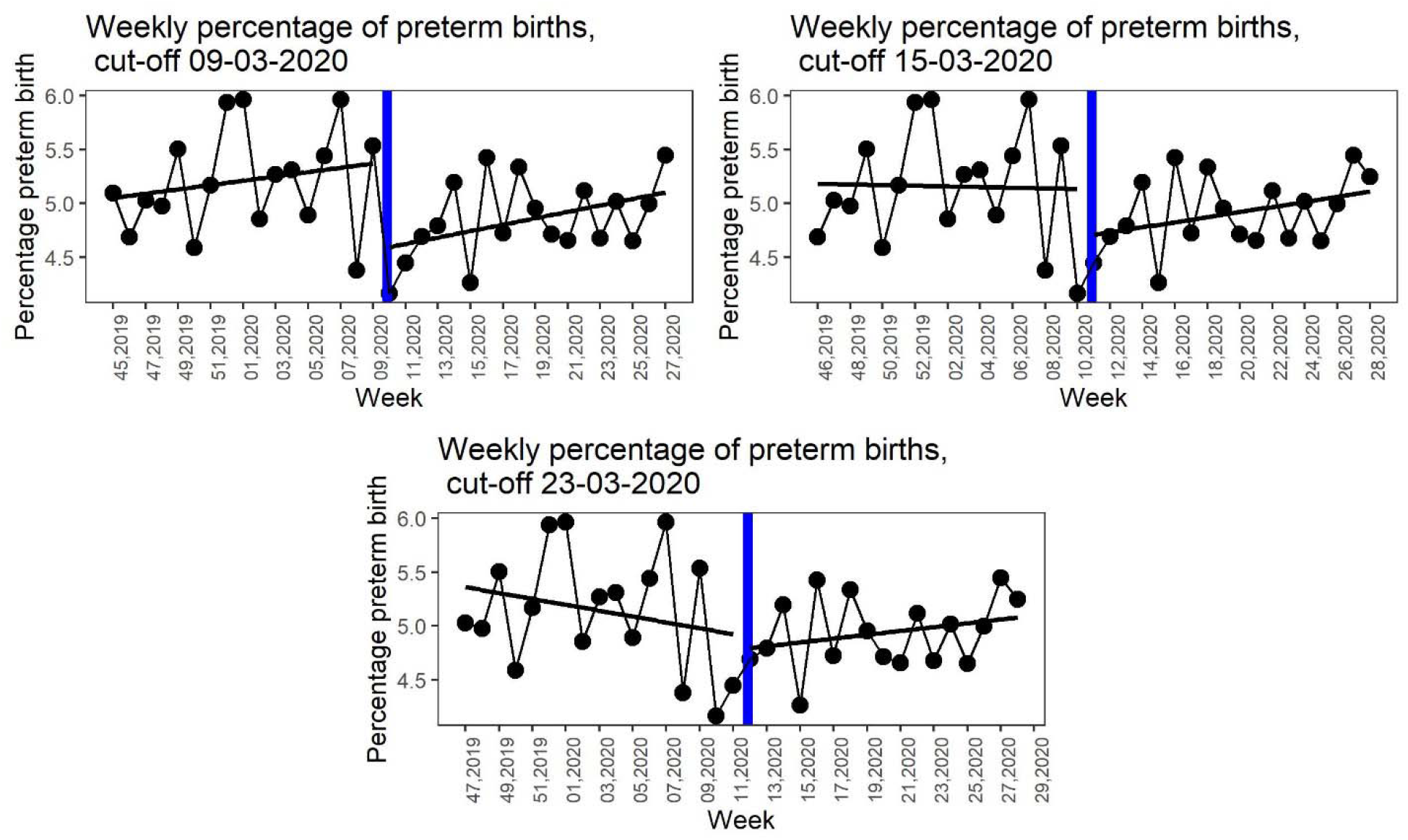
Regression discontinuity in weekly preterm birth incidence surrounding implementation of COVID-19 mitigation measures.

Given these findings and to restrict the number of analyses, we explored effect modification and conducted sensitivity analyses only for the 9 March COVID-19 mitigation measures, and only for the overall incidence of preterm birth. Although exploration of potential effect modification by neighbourhood SES suggested that the reductions in preterm birth predominantly occurred in those living in high-SES neighbourhoods, this was not statistically significant (Supplement page 15). Findings were robust to censoring of one or two weeks of data prior to or following the 9 March measures, and remained statistically significant predominantly for the two-month time window (Supplement page 16).

## Discussion

In this large national quasi-experimental study spanning a 10-year period, substantial reductions in preterm births were observed following implementation of the first national COVID-19 mitigation measures in the Netherlands. These reductions were consistent across various degrees of prematurity. No significant impact of extension of the measures introduced one and two weeks later was observed. Taken together with preliminary evidence from other countries,^2,3^ these findings open up important opportunities to help identify novel preventive strategies for preterm birth.

To the best of our knowledge, our study is by far the largest to have assessed the impact of COVID-19 mitigation measures on the incidence of preterm birth. Making use of national-level routinely collected data, we had over 1.5 million individual records available for analysis, including over 55 thousand babies born after implementation of the measures in the Netherlands. Since over 99% of babies in the Netherlands undergoes neonatal dried blood spot screening,^12^ and very few babies in the dataset had missing outcome data, our data are highly representative. By applying a quasi-experimental approach, our study progresses substantially from earlier uncontrolled before-after studies, thus maximising opportunities for causal interpretation.^19^ Our findings were in addition robust to various model specifications, further strengthening confidence in our findings.

Our study also has limitations. Given the unanticipated nature of the COVID-19 pandemic and associated mitigation measures, we had to use a retrospective approach to data collection. As in any registry-based study, there may have been registration errors, and a very small proportion of individuals had missing data. Cross-validation against Perined suggested very little temporal variation in comparability of the data or missing variables, which − even if present − should have been captured by our difference-in-regression-discontinuity design, making any impact on our effect estimation unlikely. There was a slight underrepresentation of extremely preterm and ELBW births in our dataset as compared to Perined. This was anticipated as a result of three issues: 1. aggregated birth weight data for Perined included babies born between 22+0 and 23+6 weeks, hence explaining overrepresentation of ELBW babies in Perined; 2.

Perined data include stillbirths; and 3. extremely preterm babies are at increased risk of dying in the first few days after birth.^24^ For obvious reasons, stillborn babies and those dying shortly after birth did not contribute data to the neonatal screening programme and hence were missing from our dataset. Importantly, our validation indicates that this relative underrepresentation was not differential over time and is therefore unlikely to have influenced our findings. If anything, survival of preterm babies improved over the study period, which would have biased our findings towards the null. Finally, our dataset lacked information on mode of delivery or labour induction, limiting our ability to identify any differential impact of the COVID-19 mitigation measures on spontaneous versus induced preterm births.

Our study progresses from earlier work in a number of ways, including using robust quasi-experimental methodology and having a much larger sample size.^19,20,25^ Although in the Irish study none of the January-April periods in the 19 years preceding 2020 had seen proportions of extremely-low-birth-weight (ELBW) and VLBW births as low as in 2020,^3^ the numbers of observed versus anticipated ELBW and VLWB births were very small (none versus four, and three versus 11, respectively). Furthermore, of the four months in 2020 across which births were evaluated against preceding data, only one-and-a-half were in fact post-implementation of the lockdown measures, complicating causal interpretation. Similar to ours, the Danish study used national data from the neonatal dried blood spot screening programme.^2^ Based on figures presented in their manuscript, we calculated that only one extremely preterm birth had been observed in the first month following lockdown, where five to six were expected. Again, a striking relative reduction but a small drop in absolute terms. Intriguingly, the observed reduction in preterm births in Denmark and Ireland predominantly affected the very smallest babies,^2,3^ whereas in our study the decrease was fairly constant across gestational age strata. This is important, as the vast majority of preterm babies are born moderately to late preterm (i.e. 32+0 to 36+6 weeks), and our data suggest that prevention might be possible for the smallest up to the largest groups. A comparison of birth outcomes in a London hospital before and after manifestation of the COVID-19 pandemic, revealed no changes in the incidence of preterm births or of births before 34 weeks gestation.^26^ Again, this study had a small sample size and it did not specifically investigate impact of the lockdown, hampering comparison. Interestingly, they noted an increase in stillbirths of six per 1,000 following the COVID-19 pandemic.^26^ As contemporary information on stillbirths was unavailable in our study, we could not discern whether a small part of the observed reduction in preterm births might have occurred at the expense of an increase in stillbirths.

The aetiology of spontaneous preterm birth, which accounts for roughly two-thirds of all preterm births, is largely obscure and likely multifactorial, hampering effective prevention.^27^ Many of the known risk factors for preterm birth may be positively influenced by implementation of COVID-19 mitigation measures. This includes asymptomatic maternal infection, which by means of vertical transmission can cause intrauterine infection, initiating a cascade resulting in preterm birth.^27,28^ Physical distancing and self-isolation, lack of commuting, closing of schools and childcare facilities, and increased awareness of the importance of hygiene (e.g. hand-washing) all reduce contact with pathogens, and accordingly, risk of infection. Timing of the observed preterm birth reductions directly following the first set of COVID-19 mitigation measures suggests that hygiene measures and anticipatory behavioural changes have been most instrumental. In addition, closure of most businesses and obligatory home assignments likely resulted in less physical demanding work, less shift-work, less work-related stress, optimisation of sleep duration, uptake of maternal exercise in- and outdoors, and increased social support, which all may have had a positive impact. Substantial reductions in air pollution have furthermore been reported following COVID-19 mitigation measures across the globe.^29^ Given the recognised increased risk of delivering preterm when being exposed to air pollution,^30^ this may explain part of the observed reductions. It is furthermore important to note that a large minority of preterm births is in fact iatrogenic. That is, obstetricians induce delivery, usually for maternal or fetal health concerns. As such, changes in obstetric practice or care-seeking behaviour of pregnant women may also have contributed. Finally, substantial evidence indicates that the pandemic and associated lockdown measures have aggravated existing health and socioeconomic inequalities within populations.^21,22^ In this regard, the signal in our data − albeit not statistically significant − suggesting that the reductions in preterm births were confined to people living in high-SES neighbourhoods is of considerable concern and requires further study.

Preterm birth is the primary contributor to mortality and morbidity in early childhood.^4^ Survivors are at increased risk of long-term negative consequences, including adverse cognitive and motor development,^6,7^ behavioural and mental health problems,^5,31^ and respiratory disorders.^8^ Globally, the incidence of preterm birth is on the rise,^4^ and current options for prevention are very limited.^9^ Here, we demonstrate that national introduction of COVID-19 mitigation measures in the Netherlands was associated with a 15-23% reduction in preterm births, substantiating preliminary findings from other countries.^2,3^ COVID-19 mitigation measures have been implemented across countries with substantial variation in timing, content and comprehensiveness.^32^ Similarly, levels of various risk factors for preterm delivery that might be responsive to lockdown measures also vary across populations. International collaborative efforts will be key to incorporating these sources of variation in innovative global evaluations to further study the link between COVID-19 mitigation measures and preterm births. Identification of the underlying mechanisms may then inform the development of much needed novel preventive strategies for preterm birth.

## Contributors

JVB conceived the study. JVB and LCMB developed the study protocol with involvement of EAPS and IKMR. JVB, LBO and LCMB analysed the data. All authors were involved in interpreting the data. JVB wrote the draft paper and all authors provided input at the writing stage. All authors read and approved the final version of the manuscript.

## Data Availability

The authors are open to sharing data, however consent should be sought with the Dutch National Institute for Public Health and the Environment (RIVM)

## Declaration of interests

We declare no competing interests

## Acknowledgements

We thank Roger Venema at RIVM for preparing the data extract and Martin de Vries at RIVM for facilitating data provision. No specific funding was available for this study.

## Notes

### Competing Interest Statement

The authors have declared no competing interest.

### Clinical Trial

Not applicable

### Funding Statement

No funding was received for this study

### Author Declarations

According to Dutch law (WMO) no formal ethical review was required for this study. According to standard procedures and under strict conditions that were fulfilled, RIVM allows anonymised data registered as part of the screening programme to be used for research purposes with waiver of consent. A protocol for the study was develop a priori and approved by RIVM prior to data provision.

